# Vitamin B12 deficiency in newly-arrived pregnant refugees in Calgary, Canada from 2015-2020

**DOI:** 10.1101/2025.08.24.25331053

**Authors:** Liana Hwang, Giselle DeVetten, Rachel Talavlikar, Sara Al Sharani, Mohammad Yasir Essar, Gabriel E Fabreau, Erin Hetherington

## Abstract

**Background:** Vitamin B12 deficiency is linked to adverse maternal and infant outcomes, such as preterm birth and low birth weight. Pregnant refugees are particularly vulnerable to nutritional deficiencies due to food insecurity and gastrointestinal conditions. However, limited research has explored the prevalence of B12 deficiency among pregnant refugees in Canada.

**Objective:** To determine the prevalence of vitamin B12 deficiency in pregnant refugees receiving care at the Mosaic Refugee Health Clinic (MRHC) in Calgary and to examine their demographic and clinical characteristics.

**Methods:** We conducted a retrospective chart review for pregnant patients at MRHC with estimated delivery dates between November 2015 to December 2020. We assessed the association between B12 deficiency and demographic characteristics, anemia, iron deficiency, macrocytosis, and relevant infectious diseases using chi-square tests and Fisher’s exact tests.

**Results:** Of 159 pregnant refugee patients tested for B12, 42.8% (n=68) had B12 deficiency (<220 pmol/L), 50.3% (n=80) had iron deficiency (ferritin <30 ul/L), 54.7% (n=87) were anemic (hemoglobin <110 g/L), and <5 had macrocytosis. Among patients with B12 deficiency, 58.8% (n=40) had concomitant iron deficiency. B12 deficiency was significantly associated with later gestational age at first prenatal visit (p=0.034). There was an association between B12 status and hemoglobinopathy (p=0.027), but it was not in the expected direction. No significant associations were found with other demographic or clinical characteristics (p> 0.050).

**Conclusion:** B12 and iron deficiency were common among newly arrived pregnant refugees, suggesting poor nutritional status. The absence of macrocytosis was insufficient to rule out B12 deficiency. Screening for B12 deficiency during pregnancy, while not recommended in Canada, may be considered for refugees.

## BACKGROUND

In 2022, Canada resettled 47,600 refugees, more than any other country(1). The UN recognizes refugee women are marginalized, with unique medical and psychosocial needs(2). Pregnancy is an especially vulnerable time for resettled refugees. Multiple studies indicate they have worse perinatal and neonatal outcomes(3–5).

In pregnancy, B12 deficiency may increase the risk of preterm birth and low birth weight(6). Neural tube defects are more common(7), with Molloy *et al*. suggesting B12 levels >220 pmol/L are necessary to reduce the risk of neural tube defects(7). Maternal B12 deficiency can also lead to poor feeding, infections, hypotonia and developmental regression in infants(8). Case reports have found severe B12 deficiency in infants of immigrants to Canada, and refugee infants(7,9).

There is little consensus regarding a single cutoff level for B12 deficiency. In the literature, cutoffs range from 148-220 pmol/L(10). Some guidelines recommend confirmatory testing such as methylmalonic acid levels (11), but these are considered specialty tests regionally and are rarely ordered in primary care.

B12 deficiency is uncommon in Canada, found in 4.6% of the general population (<148 pmol/L)(12). Routine screening for B12 deficiency during pregnancy is not currently recommended (13). This may reflect an assumption that most patients have adequate nutritional intake; however, multiple factors predispose refugees to nutritional deficiencies. Low animal product intake, high prevalence of *H. pylori* infection leading to gastric atrophy and malabsorption, and intestinal parasites such as *Giardia* increase the risk of B12 deficiency(14,15).

Previous studies have found high rates of B12 deficiency among resettled refugees. An Australian study revealed 16.5% of newly arrived refugees from Bhutan, Iran and Afghanistan had B12 deficiency (<150 pmol/L)(16). One study of Syrian refugee women of reproductive age in Turkey found 45.6% had B12 deficiency(17).

Relying on clinical symptoms and signs to prompt testing for B12 deficiency may be insufficient, as these are often non-specific. Pancytopenia and megaloblastic anemia may be seen, but studies have shown macrocytosis has low sensitivity for detecting B12 deficiency in newcomer populations, likely due to concomitant iron deficiency and hemoglobinopathies(16,18).

To our knowledge, there are no studies examining the prevalence of B12 deficiency in pregnant refugees. In this retrospective review, we sought to determine the prevalence of B12 deficiency among our pregnant refugees and examine their demographic characteristics and clinical findings to guide screening and treatment.

### Theoretical Framework

This study was guided by the Health Equity Framework, which examines how social and structural determinants contribute to inequities in health outcomes. The Health Equity Framework emphasizes the role of structural determinants, including immigration policies, healthcare access, and socioeconomic status, in shaping health inequities. It also considers interpersonal factors, such as language barriers and cultural differences in healthcare-seeking behavior, that may influence access to nutritional screening.

Refugee populations face unique vulnerabilities due to factors such as forced displacement, limited access to healthcare, food insecurity, and socioeconomic barriers, all of which can impact nutritional status and maternal health. By applying this framework, we aimed to explore how refugee status may contribute to inequities in vitamin B12 deficiency, anemia, iron deficiency, macrocytosis, and infectious diseases during pregnancy.

## METHODS

### Participants and Data Collection

The Mosaic Refugee Health Clinic (MRHC) in Calgary is a specialized interdisciplinary clinic providing medical services, including prenatal care, to refugees and refugee claimants within the first two years of arrival in Canada. Government-assisted refugees are connected with the clinic by resettlement agencies, while privately sponsored refugees and refugee claimants can self-refer.

Patients undergo an intake visit, during which demographic characteristics are collected and entered into a secure electronic medical record. Routine screening lab investigations are ordered, including CBC, ferritin, hemoglobinopathy screen, and serum B12. Serological testing for *Schistosoma* and *Strongyloides* is conducted based on patient’s country of origin. *H. pylori* testing and stool testing for ova and parasites (O&P) are performed based on clinical symptoms and signs.

During pregnancy, routine lab investigations are ordered at the first prenatal visit including CBC, ferritin, serum B12 and hemoglobinopathy screen if not already done at intake. Patients are followed by family doctors, in collaboration with obstetricians if their pregnancies are high risk. A small number of patients are transferred outside of the clinic due to specialized care needs.

A total of 185 patients who received prenatal care at MRHC with due dates between November 2015 to December 2020 were included. Two twin pregnancies were excluded. Of the remaining 183 patients, 7 had no recorded B12 levels, and 17 did not have B12 measured during pregnancy, leaving 159 patients in the final analysis.

Data were extracted from the EMR at MRHC via manual chart review and electronic extraction. We included the following demographic variables: region of origin, year of arrival, language, need for interpreter, refugee type, maternal age, gravidity, and gestational age at first prenatal visit. We collected the following lab data: serum B12, Hb, ferritin, MCV, hemoglobinopathy screen results, as well as *H. pylori* test results, serology for *Schistosomiasis* and *Strongyloides*, and stool ova and parasites (O&P). All automated searches were cross-checked manually to ensure validity.

At the time, guidelines did not recommend screening asymptomatic patients for *H. pylori* (19,20) and O&P (21). Therefore, when *H. pylori* and O&P were not ordered, we considered this equivalent to a negative result. In routine practice, the clinic does not test for H. pylori or intestinal parasites during pregnancy and defers treatment until after delivery. Thus, we included any *H. pylori* and intestinal parasite results that had been done during the patient’s time at the clinic, generally a period of two years or less.

For all other lab values (hemoglobin, ferritin and MCV), complete data was included.

## Measures

### B12 Deficiency

Vitamin B12 deficiency was defined as serum B12 level <220 pmol/L, the threshold identified by Molloy *et al*. as necessary to prevent neural tube defects. (7)

### Anemia, Iron Deficiency, Macrocytosis and Hemoglobinopathies

Anemia was defined according to the World Health Organization (WHO) criteria for pregnancy as hemoglobin levels <110 g/L (22). Iron deficiency was defined as ferritin level <30 µg/L. Macrocytosis, which can be a marker of B12 or folate deficiency, was defined as a mean corpuscular volume (MCV) >100 fL, consistent with the local laboratory reference range (23)

For patients with multiple B12, hemoglobin, ferritin, and MCV measurements during a single pregnancy, only the first value was used in the analysis to minimize potential confounding from treatment interventions.

In the clinic’s health region, hemoglobin electrophoresis is performed as the initial test for screening for hemoglobinopathies, followed by reflex molecular testing when warranted.

### Testing for *H. pylori* and Intestinal Parsites

Testing for *H*. pylori and intestinal parasites was conducted based on regional laboratory testing protocols. In January 2018, the regional laboratory transitioned from the urea breath test to the stool antigen test for diagnosis of *H. pylori* infection(24).

The regional laboratory employs a polymerase chain reaction screen method which detects *Giardia, Cryptosporidium* and *Entamoeba histolytica*(25). Full O&P testing is performed only when clinically indicated.

### Analysis

A retrospective review of demographic and clinical characteristics was performed.

Demographic characteristics of categorical data (region of origin, year of arrival, language spoken, need for interpreter, refugee type, and parity) are described using counts and percentages. Maternal characteristics of continuous variables (age at delivery, gravidity, and gestational age at first visit) are summarized as means with standard deviations. The prevalence of B12 deficiency and other comorbidities are described as counts and percentages.

We compared demographic characteristics of patients with and without B12 deficiency using chi-square tests or t-tests as appropriate. For categorical comparisons with expected cell counts less than 5, we used Fischer’s exact tests.

We examined concomitant conditions with B12 deficiency using chi-square tests and Fischer’s exact tests based on cell size.

Cell sizes less than 5 were suppressed to protect patient confidentiality.

## RESULTS

Patient demographic and clinical characteristics are shown in Table 1a and 1b. The top regions of origin were Africa (86 patients, 54.1%) and the Middle East (48 patients, 30.2%), with the top four countries of origin being Eritrea, Syria, Ethiopia and Iraq. The most commonly spoken languages were Tigrinya (44, patients 27.7%) and Arabic (44 patients, 27.7%). The mean maternal age was 29.9 years (SD= 5.6). More than 80% of patients were multiparous, with a mean gravidity of 3.2 (SD= 1.7). Most patients presented early in pregnancy, with a mean gestational age at first prenatal visit of 14.7 weeks (SD= 9.7).

**Table 1a:**
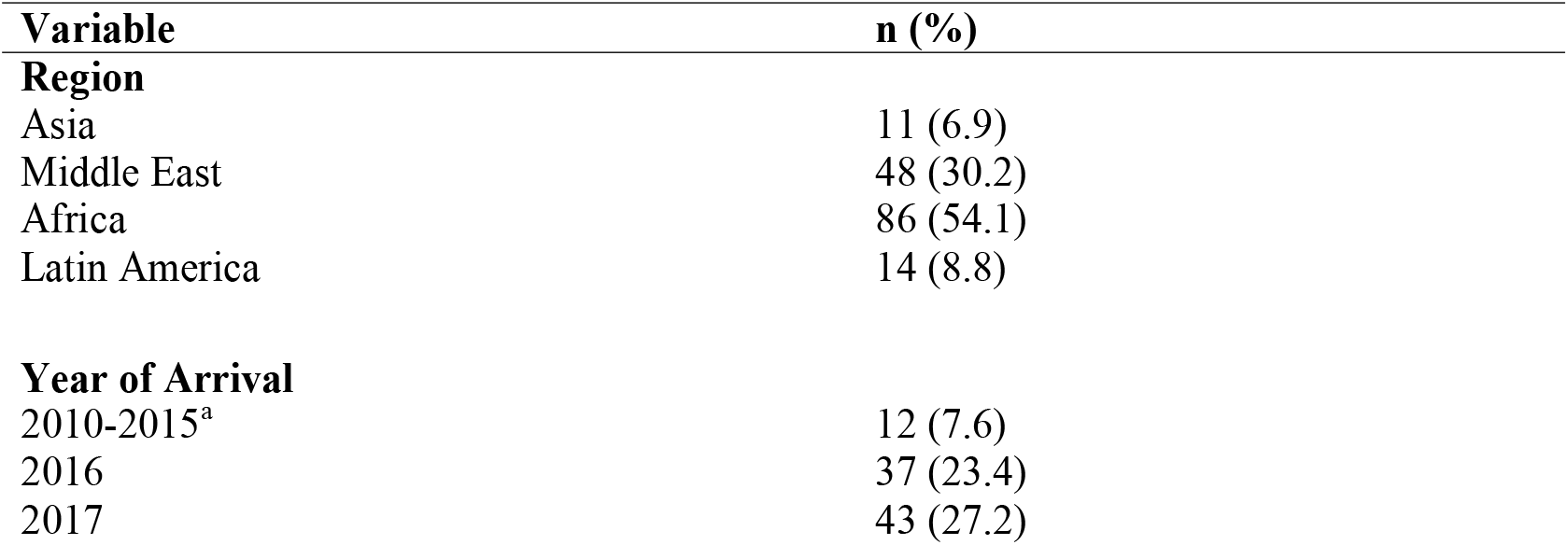
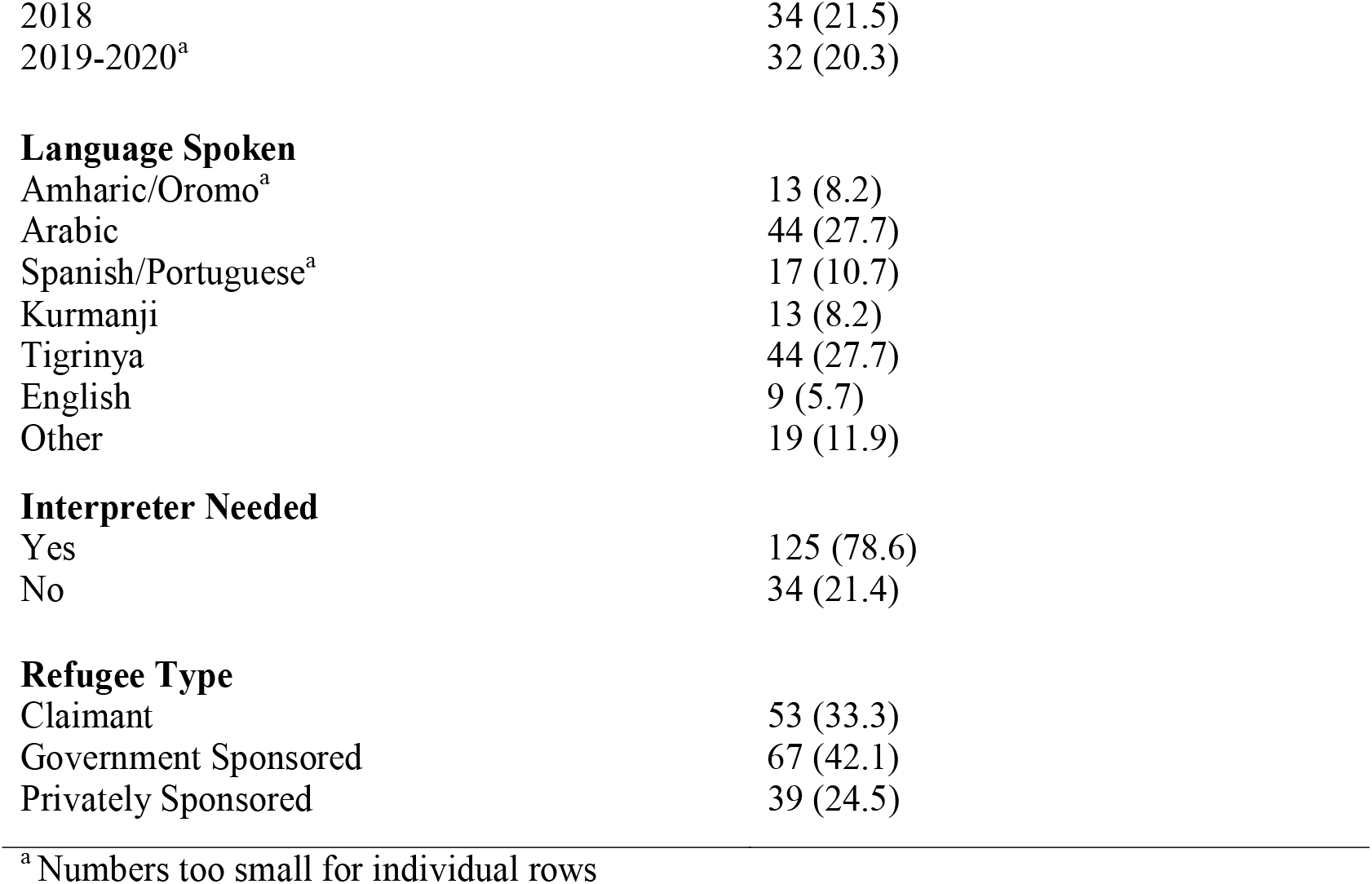
Study cohort characteristics.

| Variable | n (%) |
| --- | --- |
| **Region** |  |
| Asia | 11 (6.9) |
| Middle East | 48 (30.2) |
| Africa | 86 (54.1) |
| Latin America | 14 (8.8) |
| **Year of Arrival** |  |
| 2010-2015<sup>a</sup> | 12 (7.6) |
| 2016 | 37 (23.4) |
| 2017 | 43 (27.2) |
| 2018 | 34 (21.5) |
| 2019-2020<sup>a</sup> | 32 (20.3) |
| Language Spoken |  |
| Amharic/Oromo<sup>a</sup> | 13 (8.2) |
| Arabic | 44 (27.7) |
| Spanish/Portuguese<sup>a</sup> | 17 (10.7) |
| Kurmanji | 13 (8.2) |
| Tigrinya | 44 (27.7) |
| English | 9 (5.7) |
| Other | 19 (11.9) |
| Interpreter Needed |  |
| Yes | 125 (78.6) |
| No | 34 (21.4) |
| Refugee Type |  |
| Claimant | 53 (33.3) |
| Government Sponsored | 67 (42.1) |
| Privately Sponsored | 39 (24.5) |

<sup>a</sup> Numbers too small for individual rows

**Table 1b:**
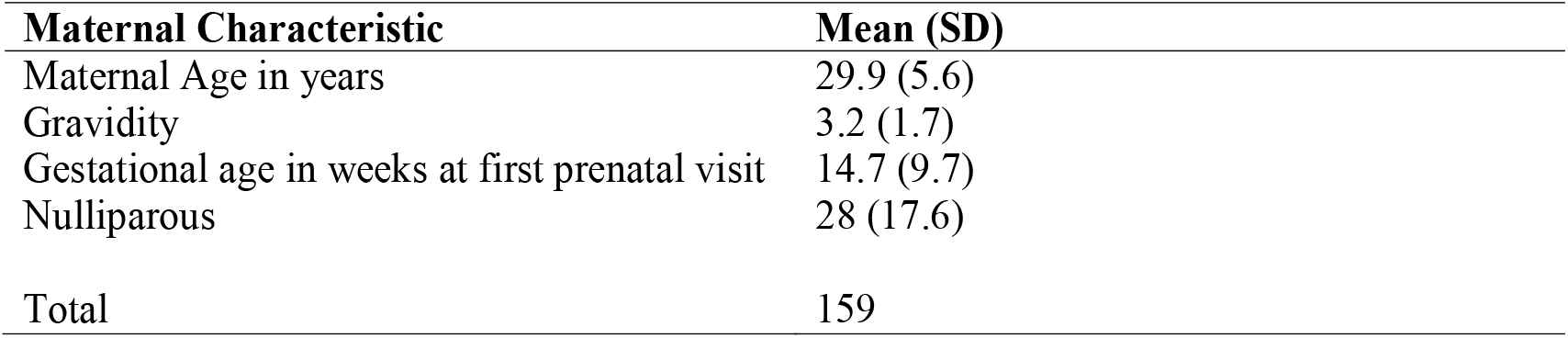
Maternal clinical and demographic characteristics.

B12 deficiency was common, affecting 68 patients (42.8%) (Table 2). Of these, 50% had their B12 measured in the first trimester, 28% in the second trimester, and 22% in the third trimester. Anemia and iron deficiency were also highly prevalent, with 87 patients (54.7%) meeting the criteria for anemia (Hb <110 g/L) and 80 patients (50.3%) classified as iron deficient (ferritin < 30 ug/L). Macrocytosis was rare, exhibited by fewer than five patients (Table 2). A positive hemoglobinopathy screen was identified in 17 patients (10.7%).

**Table 2:**
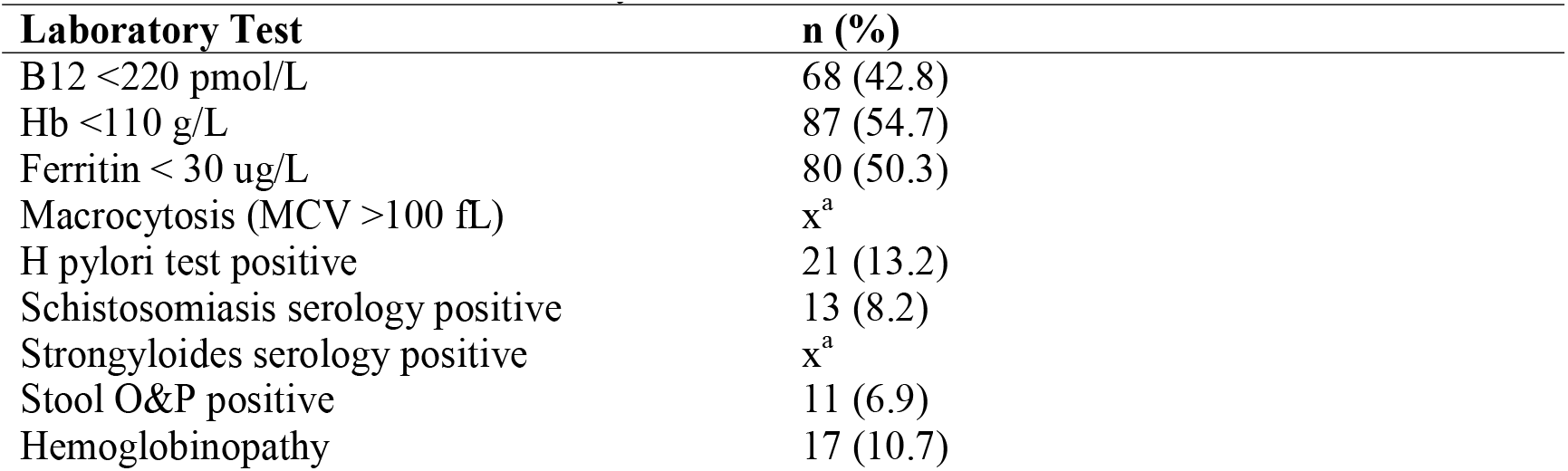
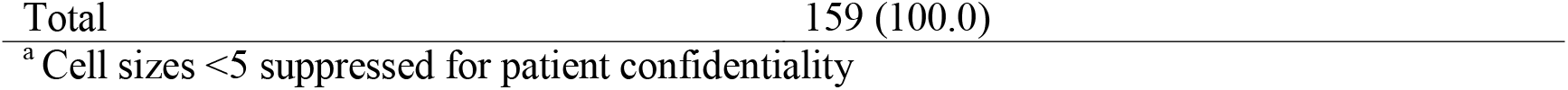
Prevalence of B12 deficiency and other comorbidities.

| Laboratory Test | n (%) |
| --- | --- |
| B12 <220 pmol/L | 68 (42.8) |
| Hb <110 g/L | 87 (54.7) |
| Ferritin < 30 ug/L | 80 (50.3) |
| Macrocytosis (MCV >100 fL) | x<sup>a</sup> |
| H pylori test positive | 21 (13.2) |
| Schistosomiasis serology positive | 13 (8.2) |
| Strongyloides serology positive | x<sup>a</sup> |
| Stool O&P positive | 11 (6.9) |
| Hemoglobinopathy | 17 (10.7) |

<sup>a</sup> Cell sizes <5 suppressed for patient confidentiality

H. pylori infection was identified in 21 patients (13.2%). *Schistosoma* serology was positive in 13 patients (8.2%). Intestinal parasites were identified in 11 patients (6.9%) (Table 2). The most commonly identified intestinal parasites included *Blastocystis hominis, Giardia lamblia, Hymenolepis nana, Dientamoeba fragilis*, and hookworm. Fewer than five patients tested positive for *Strongyloides*.

B12 deficiency was not significantly associated with any maternal characteristics (p> 0.050) (Table 3a) except for gestational age at first visit (p= 0.034). Patients with B12 deficiency presented for prenatal care approximately a month later than patients without B12 deficiency (p=0.03). For coexisting conditions, a significant association was observed between B12 deficiency status and a negative hemoglobinopathy screen (p = 0.027, Table 3b).

**Table 3a:**
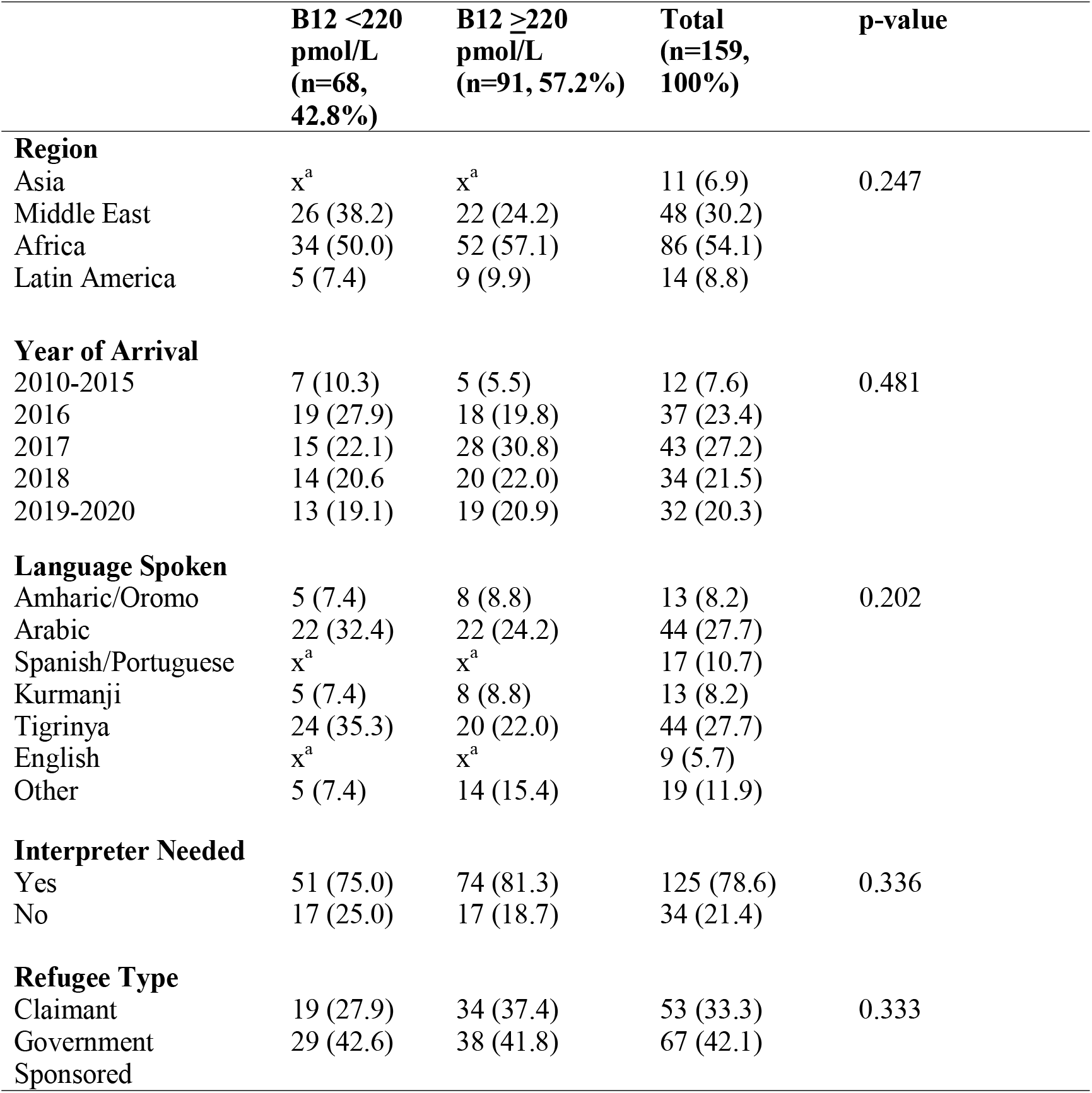
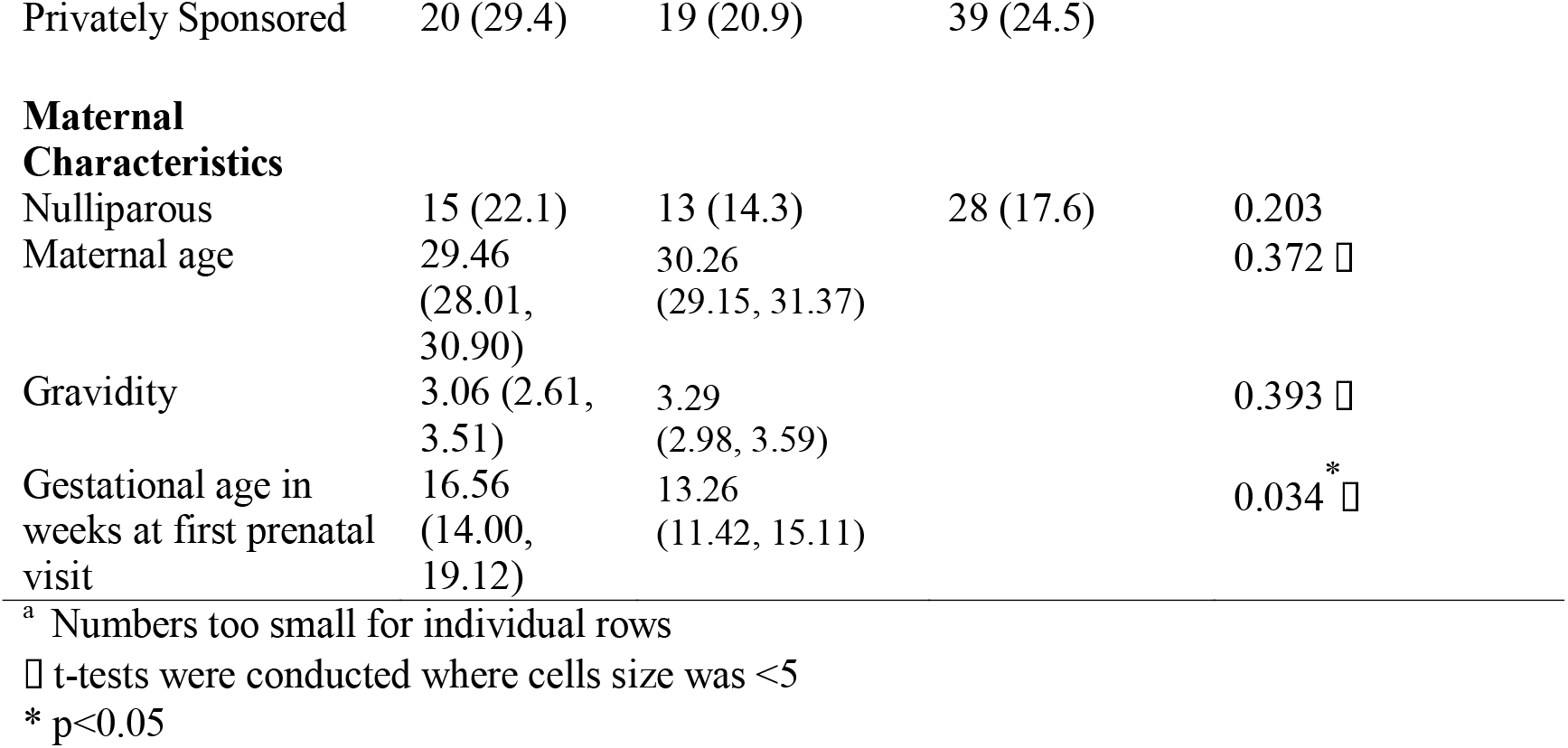
Demographic Characteristics by B12 status.

**Table 3b:**
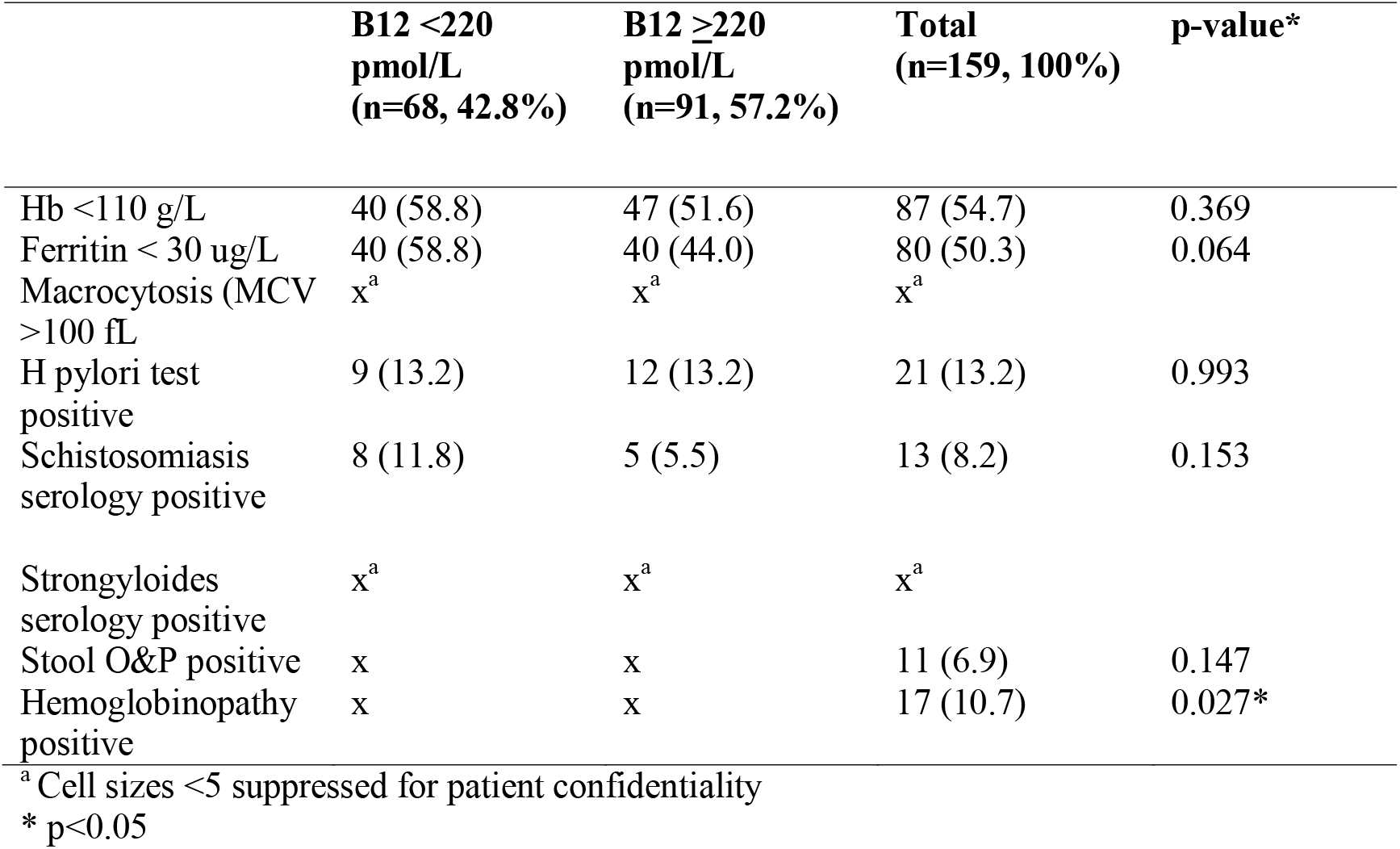
Concomitant conditions by B12 status.

|  | **B12 <220**<br>**pmol/L**<br>**(n=68, 42.8%)** | **B12 ≥220**<br>**pmol/L**<br>**(n=91, 57.2%)** | **Total**<br>**(n=159, 100%)** | **p-value\*** |
| --- | --- | --- | --- | --- |
| Hb <110 g/L | 40 (58.8) | 47 (51.6) | 87 (54.7) | 0.369 |
| Ferritin < 30 ug/L | 40 (58.8) | 40 (44.0) | 80 (50.3) | 0.064 |
| Macrocytosis (MCV >100 fL) | x<sup>a</sup> | x<sup>a</sup> | x<sup>a</sup> |  |
| H pylori test positive | 9 (13.2) | 12 (13.2) | 21 (13.2) | 0.993 |
| Schistosomiasis serology positive | 8 (11.8) | 5 (5.5) | 13 (8.2) | 0.153 |
| Strongyloides serology positive | x<sup>a</sup> | x<sup>a</sup> | x<sup>a</sup> |  |
| Stool O&P positive | x | x | 11 (6.9) | 0.147 |
| Hemoglobinopathy positive | x | x | 17 (10.7) | 0.027* |

<sup>a</sup> Cell sizes <5 suppressed for patient confidentiality
\* p<0.05

## DISCUSSION

B12 deficiency in pregnancy can lead to severe maternal and newborn outcomes, but routine screening is not currently recommended in Canadian-born populations (26). B12 deficiency was common among recently arrived pregnant refugees in Calgary, Canada. Nearly half had B12 deficiency, defined as <220 pmol/L, the threshold necessary to reduce the risk of neural tube defects. We were unable to identify any characteristics which could allow us to selectively screen a subset of our population for B12 deficiency.

More than half of our cohort had iron deficiency, nearly double the rate reported among a cohort of 532 female patients with a median age of 31.2 years receiving care at MRHC from 2011-2015(27). One in ten had a hemoglobinopathy; both iron deficiency and hemoglobinopathies are associated with microcytosis and may mask macrocytosis. Further, a negligible number of patients in our study had macrocytosis; thus, relying on macrocytosis to prompt B12 testing would have been inadequate in our population.

*H. pylori* and intestinal parasites were common in our population and have been identified as risk factors for B12 deficiency(14,15), but there was no association in our population. Previous research has found hemoglobinopathies are associated with B12 deficiency due to increased erythropoiesis(28), but our study found the opposite. These findings should be interpreted with caution due to our small sample size.

Later gestational age at first prenatal visit was the only maternal characteristic associated with B12 deficiency and may be related to the fact that B12 levels fall throughout pregnancy, decreasing by up to 30% by the third trimester due to dilution, fetal uptake, hormonal changes, and other mechanisms which have not yet been elucidated(29,30). Pregnancy-specific and trimester-specific B12 cutoffs have not been established and would merit further research.

As it stands, our results resemble those of other Canadian studies, which found 31-52% of patients had B12 levels <220 pmol/L in early pregnancy(29,31). MRHC’s practice is to test for B12 deficiency at intake, meaning that for many patients, B12 deficiency is identified and treated pre-conception. Without intake screening, the prevalence of B12 deficiency may have been even higher.

Our study adds to the existing literature as our cohort was mainly from Africa and the Middle East, while previous Canadian studies have mainly included white and Asian women(29,31,32).

B12 deficiency in pregnancy can have devastating fetal, neonatal and infant consequences but is relatively simple to treat. Both oral and IM formulations of B12 are recommended, depending on the severity of the deficiency and gestational age of the patient. High-dose oral B12 is tolerated well, with minimal side effects or risks(33). Currently, the Society of Obstetricians and Gynecologists of Canada only recommends the small amount of B12 in a prenatal vitamin during pregnancy(34), which is inadequate for treatment of B12 deficiency.

Our study confirms B12 deficiency is common among recently arrived pregnant refugees. Screening for and treating B12 deficiency in pregnant refugees is likely beneficial, carries minimal risk and potentially improves equity in care to these vulnerable patients.

Preventive supplementation may be indicated; for example, the World Health Organization recommends consideration of universal iron supplementation instead of screening in populations with a severe prevalence of anemia (>40%)(35). Further research is needed to clarify the optimal approach to B12 deficiency during pregnancy.

### Limitations

Our results are limited by our small sample size. We may have been underpowered to assess certain outcomes. In particular, it’s possible there is an association between iron deficiency and B12 deficiency since both are related to poor nutritional intake.

However, one of our study strengths is that the clinic and thus, our cohort, captures an entire geographic area. Calgary currently resettles among the highest number of refugees per capita in Canada(36), and refugee health is highly centralized in Calgary. Additionally, most of MRHC’s pregnant patients receive their prenatal care at MRHC rather than being referred elsewhere.

### New Contribution to the Literature

Our study contributes to the growing body of literature on B12 deficiency during pregnancy by presenting data specific to the refugee population. Our participants, primarily from the Middle East and Africa, offer novel insights into the prevalence and clinical characteristics of B12 deficiency in these understudied populations.

Nearly half of our participants had B12 levels which fell below the level required to reduce the risk of neural tube defects. Unlike previous research which has linked B12 deficiency to specific risk factors, such as *H. pylori* infection and intestinal parasites, our study did not find these associations, suggesting the need for further investigation into the mechanisms of B12 deficiency among refugees. Given the ease of treatment and potential impact on maternal and neonatal outcomes, our findings contribute to the discussion on whether targeted screening or preventative supplementation should be considered for pregnant refugees.

## Data Availability

All data produced in the present study are available upon reasonable request to the authors

## REFERENCES

1. UNHCR [Internet]. [cited 2024 Oct 20]. Global Trends Report 2022. Available from: https://www.unhcr.org/global-trends-report-2022

2. Refworld [Internet]. [cited 2024 Oct 20]. New York Declaration for Refugees and Migrants□: resolution /adopted by the General Assembly. Available from: https://www.refworld.org/legal/resolution/unga/2016/en/112142

3. Kandasamy T, Cherniak R, Shah R, Yudin MH, Spitzer R. Obstetric Risks and Outcomes of Refugee Women at a Single Centre in Toronto. Journal of Obstetrics and Gynaecology Canada. 2014 Apr;36(4):296–302.

4. Johnson EB, Reed SD, Hitti J, Batra M. Increased risk of adverse pregnancy outcome among Somali immigrants in Washington state. American Journal of Obstetrics and Gynecology. 2005 Aug;193(2):475–82.

5. Gibson-Helm M, Teede H, Block A, Knight M, East C, Wallace EM, et al. Maternal health and pregnancy outcomes among women of refugee background from African countries: a retrospective, observational study in Australia. BMC Pregnancy Childbirth. 2014 Dec;14(1):392.

6. Rogne T, Tielemans MJ, Chong MFF, Yajnik CS, Krishnaveni GV, Poston L, et al. Associations of Maternal Vitamin B12 Concentration in Pregnancy With the Risks of Preterm Birth and Low Birth Weight: A Systematic Review and Meta-Analysis of Individual Participant Data. Am J Epidemiol. 2017 Jan 20;amjepid;kww212v1.

7. Molloy AM, Kirke PN, Troendle JF, Burke H, Sutton M, Brody LC, et al. Maternal Vitamin B12 Status and Risk of Neural Tube Defects in a Population With High Neural Tube Defect Prevalence and No Folic Acid Fortification. Pediatrics. 2009 Mar 1;123(3):917–23.

8. Roumeliotis N, Dix D, Lipson A. Vitamin B _12_ deficiency in infants secondary to maternal causes. CMAJ. 2012 Oct 2;184(14):1593–8.

9. Belen B, Hismi BO, Kocak U. Severe vitamin B _12_ deficiency with pancytopenia, hepatosplenomegaly and leukoerythroblastosis in two Syrian refugee infants: a challenge to differentiate from acute leukaemia. BMJ Case Reports. 2014 Mar 5;bcr2014203742.

10. Allen LH. How common is vitamin B-12 deficiency? The American Journal of Clinical Nutrition. 2009 Feb;89(2):693S–696S.

11. Langan RC, Goodbred AJ. Vitamin B12 Deficiency: Recognition and Management. Am Fam Physician. 2017 Sep 15;96(6):384–9.

12. MacFarlane AJ, Greene-Finestone LS, Shi Y. Vitamin B-12 and homocysteine status in a folate-replete population: results from the Canadian Health Measures Survey. The American Journal of Clinical Nutrition. 2011 Oct;94(4):1079–87.

13. Canada PHA of. Care during pregnancy: Family-centred maternity and newborn care national guidelines [Internet]. 2021 [cited 2024 Oct 20]. Available from: https://www.canada.ca/en/public-health/services/publications/healthy-living/maternity-newborn-care-guidelines-chapter-3.html

14. Kaptan K, Beyan C, Ural AU, Çetin T, Avcu F, Gülşen M, et al. Helicobacter pylori—Is It a Novel Causative Agent in Vitamin B12 Deficiency? Arch Intern Med. 2000 May 8;160(9):1349.

15. Stabler SP, Allen RH. VITAMIN B12 DEFICIENCY AS A WORLDWIDE PROBLEM. Annu Rev Nutr. 2004 Jul 14;24(1):299–326.

16. Benson J, Phillips C, Kay M, Webber MT, Ratcliff AJ, Correa-Velez I, et al. Low Vitamin B12 Levels among Newly-Arrived Refugees from Bhutan, Iran and Afghanistan: A Multicentre Australian Study. Wang G, editor. PLoS ONE. 2013 Feb 28;8(2):e57998.

17. Şimşek Z, Yentur Doni N, Gül Hilali N, Yildirimkaya G. A community-based survey on Syrian refugee women’s health and its predictors in Şanliurfa, Turkey. Women & Health. 2018 Jul 3;58(6):617–31.

18. Gupta AK, Damji A, Uppaluri A. Vitamin B12 deficiency. Prevalence among South Asians at a Toronto clinic. Can Fam Physician. 2004 May;50:743–7.

19. Helicobacter pylori | CDC Yellow Book 2024 [Internet]. [cited 2024 Oct 20]. Available from: https://wwwnc.cdc.gov/travel/yellowbook/2024/infections-diseases/helicobacter-pylori

20. Chey WD, Leontiadis GI, Howden CW, Moss SF. ACG Clinical Guideline: Treatment of Helicobacter pylori Infection. American Journal of Gastroenterology. 2017 Feb;112(2):212– 39.

21. Pottie K, Greenaway C, Feightner J, Welch V, Swinkels H, Rashid M, et al. Evidence-based clinical guidelines for immigrants and refugees. Canadian Medical Association Journal. 2011 Sep 6;183(12):E824–925.

22. Organization WH. Haemoglobin concentrations for the diagnosis of anaemia and assessment of severity. Concentrations en hémoglobine permettant de diagnostiquer l’anémie et d’en évaluer la sévérité [Internet]. 2011 [cited 2024 Oct 20]; Available from: https://iris.who.int/handle/10665/85839

23. CBC and Differential Provincial Reference Intervals and Critical Values [Internet]. Alberta Precision Laboratories; [cited 2025 Jan 15]. Available from: https://www.albertahealthservices.ca/assets/wf/lab/if-lab-cbc-and-differential-provincial-reference-intervals-and-critical-values-epic.pdf

24. Ma I, Guo M, Pillai DR, Church DL, Naugler C. Is the Utilization of Helicobacter pylori Stool Antigen Tests Appropriate in an Urban Canadian Population? American Journal of Clinical Pathology. 2020 Apr 15;153(5):686–94.

25. Transition of Testing for Stool for Ova & Parasites [Internet]. Available from: https://www.albertahealthservices.ca/assets/wf/lab/wf-lab-cls-memo-transition-of-testing-for-stool-for-ova-and-parasites.pdf

26. O’Connor DL, Blake J, Bell R, Bowen A, Callum J, Fenton S, et al. Canadian Consensus on Female Nutrition: Adolescence, Reproduction, Menopause, and Beyond. Journal of Obstetrics and Gynaecology Canada. 2016 Jun;38(6):508-554.e18.

27. Davidson MB, Brown G, Street L, McBrien K, Norrie E, Hull A, et al. Iron deficiency, anemia and association with refugee camp exposure among recently resettled refugees: A Canadian retrospective cohort study. Aoun M, editor. PLoS ONE. 2022 Dec 15;17(12):e0278838.

28. Andreadis P, Theodoridou S, Pasakiotou M, Arapoglou S, Gigi E, Vetsiou E, et al. Vitamin B12 Deficiency and Hemoglobin H Disease Early Misdiagnosed as Thrombotic Thrombocytopenic Purpura: A Series of Unfortunate Events. Case Reports in Hematology. 2015;2015:1–5.

29. Visentin CE, Masih SP, Plumptre L, Schroder TH, Sohn KJ, Ly A, et al. Low Serum Vitamin B-12 Concentrations Are Prevalent in a Cohort of Pregnant Canadian Women. The Journal of Nutrition. 2016 May;146(5):1035–42.

30. Devalia V, Hamilton MS, Molloy AM, the British Committee for Standards in Haematology. Guidelines for the diagnosis and treatment of cobalamin and folate disorders. Br J Haematol. 2014 Aug;166(4):496–513.

31. Wu BT, Innis SM, Mulder KA, Dyer RA, King DJ. Low plasma vitamin B-12 is associated with a lower pregnancy-associated rise in plasma free choline in Canadian pregnant women and lower postnatal growth rates in their male infants. The American Journal of Clinical Nutrition. 2013 Nov;98(5):1209–17.

32. Jeruszka-Bielak M, Isman C, Schroder T, Li W, Green T, Lamers Y. South Asian Ethnicity Is Related to the Highest Risk of Vitamin B12 Deficiency in Pregnant Canadian Women. Nutrients. 2017 Mar 23;9(4):317.

33. Vidal-Alaball J, Butler CC, Cannings-John R, Goringe A, Hood K, McCaddon A, et al. Oral vitamin B12 versus intramuscular vitamin B12 for vitamin B12 deficiency. Cochrane Database Syst Rev. 2005 Jul 20;(3):CD004655.

34. Wilson RD, O’Connor DL. Guideline No. 427: Folic Acid and Multivitamin Supplementation for Prevention of Folic Acid–Sensitive Congenital Anomalies. Journal of Obstetrics and Gynaecology Canada. 2022 Jun;44(6):707-719.e1.

35. Iron Deficiency Anaemia Assessment, Prevention and Control: A guide for programme managers. World Health Organization;

36. Canadian Refugee Healthcare System Atlas [Internet]. Refugee Health YYC; [cited 2025 Jan 15]. Available from: https://rh2c.org/atlas

